# Persistent symptoms after Covid-19: qualitative study of 114 “long Covid” patients and draft quality criteria for services

**DOI:** 10.1101/2020.10.13.20211854

**Authors:** Emma Ladds, Alex Rushforth, Sietse Wieringa, Sharon Taylor, Clare Rayner, Laiba Husain, Trisha Greenhalgh

**Author notes:** Corresponding author: T Greenhalgh, Nuffield Department of Primary Care Health Sciences, University of Oxford, Oxford OX2 6GG, UK.

## Abstract

**Background:** Approximately 10% of patients with Covid-19 experience symptoms beyond 3-4 weeks. Patients call this “long Covid”. We sought to document the lived experience of such patients, their accounts of accessing and receiving healthcare, and their ideas for improving services.

**Method:** We held 55 individual interviews and 8 focus groups (n = 59) with people recruited from UK-based long Covid patient support groups, social media and snowballing. We restricted some focus groups to health professionals since they had already self-organised into online communities. Participants were invited to tell their personal stories and comment on others’ stories. Data were audiotaped, transcribed, anonymised and coded using NVIVO. Analysis incorporated sociological theories of illness, healing, peer support, the clinical relationship, access to care, and service redesign.

**Results:** The sample was 70% female, aged 27-73 years, and comprised White British (74%), Asian (11%), White Other (7%), Black (4%), and Mixed (4%). 27 were doctors and 23 other health professionals. 10% had been hospitalised. Analysis revealed a confusing illness with many, varied and often relapsing-remitting symptoms and uncertain prognosis; a heavy sense of loss and stigma; difficulty accessing and navigating services; difficulty being taken seriously and achieving a diagnosis; disjointed and siloed care (including inability to access specialist services); variation in standards (e.g. inconsistent criteria for seeing, investigating and referring patients); variable quality of the therapeutic relationship (some participants felt well supported while others felt “fobbed off”); and possible critical events (e.g. deterioration after being unable to access services). Emotional touch points in participants’ experiences informed ideas for improving services.

**Conclusion:** Quality principles for a long Covid service should include ensuring access to care, reducing burden of illness, taking clinical responsibility and providing continuity of care, multi-disciplinary rehabilitation, evidence-based investigation and management, and further development of the knowledge base and clinical services.

**Study registration:** NCT04435041

## Background

“Long” Covid is the name patients gave to Covid-19 infection that has not got better yet.^1^ Working definitions of ‘post-acute’ (symptoms beyond 3-4 weeks) and ‘chronic’ (symptoms beyond 12 weeks) Covid-19 are yet to be formally confirmed.^2 3^ A positive test for Covid-19 is not a prerequisite for diagnosis of post-acute or chronic disease, since many people were never tested.^4^

The prevalence and patterning of persistent symptoms after Covid-19 is contested.^5^ Mainstream medical opinion considers them commoner in people with conditions such as asthma, diabetes and autoimmune disorders (though they are also known to occur in those with no pre-existing conditions) ^4 6-8^ and in those who were admitted to hospital.^8-10^ However, there has been little or no systematic research on people who were not hospitalised and it is even conceivable that a protracted illness may be *more* common in those whose acute illness was less severe. Whilst academic publications have estimated that 10-20% of people are still unwell after 3 weeks and 1-3% are still significantly unwell after 12 weeks,^3 8^ self-surveys of patients recruited from long Covid peer support groups suggest a much high incidence of persistent symptoms even taking account of sampling bias (for example, several thousand people from the UK in such groups report symptoms six months after their acute illness, which suggests that the figure of 1% cannot be correct).^11 12^ The high proportion of women in long Covid support groups^4 12^ may or may not reflect a true gender difference in incidence.

People with long Covid experience a confusing array of persistent and fluctuating symptoms including cough, breathlessness, fever, sore throat, chest pain, palpitations, cognitive deficits, myalgia, neurological symptoms, skin rashes, and diarrhoea; some also have persistent or intermittent low oxygen saturations.^2 4 9-13^ The cause of persisting symptoms is unknown, but probably involves several different disease mechanisms including an inflammatory reaction with a vasculitic component.^14-16^ Documented post-acute sequelae include myo- or pericarditis, heart failure, arrhythmias, and thrombo-embolic complications including myocardial infarction, stroke and venous thrombosis.^17 18^

Persisting symptoms seem to fall into three broad patterns:^4 11^ people who were initially hospitalised with acute respiratory distress syndrome (ARDS) and now have long-term respiratory symptoms dominated by breathlessness; people who were (usually) not hospitalised initially but who now have a multisystem disease with evidence of cardiac, respiratory, or neurological end-organ damage manifesting in a variety of ways; and people who have persisting symptoms, often but not always dominated by fatigue, with no evidence of organ damage. The second two groups have been little studied, and – surprisingly – were excluded from a recently-established long-term follow-up study of Covid-19 (which is limited to those who were hospitalised).^19^

Despite preliminary guidance from multiple sources,^2 20-24^ there is not yet a consistent approach to the diagnosis, management and follow-up of patients with long Covid. In the UK, the National Institute for Health and Clinical Excellence, Scottish Intercollegiate Guidelines Network, and Royal College of General Practitioners are collaborating on a more definitive guideline and NHS England has allocated funding for a new Long Covid service.^25^ To be effective, all these initiatives need to be informed not only by objective research evidence of the accuracy of tests and efficacy of treatments, but also by research on the *subjective* evidence of the patient experience.^26^

In this study, we sought to answer three key questions. First, how do people with long Covid (including those who were never hospitalised) experience the development, course and perhaps resolution of the illness over time? Second, what services have they accessed (or tried to access), and what was their experience of those services? Third, what are their ideas for improving the management of their condition and the design and delivery of services?

## Methods

### Management and governance

The dataset reported here emerged from two research studies: a mixed-method study of how patients and staff experienced remote care in the Covid-19 pandemic (funded by UK Research and Innovation Covid-19 Emergency Fund), and a funded extension, designed to embrace Covid-19, of an ongoing qualitative study of the challenges of living with multiple health conditions (from the Wellcome Trust). The work received ethical approval from the East Midlands – Leicester Central Research Ethics Committee (IRAS Project ID: 283196; REC ref 20/EM0128) on 4^th^ May 2020 and subsequent amendments. It was overseen by an independent advisory group with patient representation and a lay chair which met 3-monthly via video link. People with lived experience of Covid-19 helped interpret and analyse the data and gave feedback on drafts of the paper.

### Study design and setting

Participants were invited to choose between an individual narrative interview or participation in an online focus group. The study was conducted in UK between May and September 2020 (towards the end of the first wave of the pandemic).

### Sampling and recruitment

We originally sought to interview people who had had telephone or video consultations for acute Covid-19, as part of a wider study of remote services. We identified those individuals first via Twitter, and the link was shared on a then nascent Facebook support group (https://www.facebook.com/groups/longcovid). This brought a large response from interviewees reporting symptoms many weeks after their acute illness. We decided to undertake a focused study of such people with the following inclusion criteria: symptoms developed between February and July 2020 following an acute illness consistent with Covid-19; symptoms continued beyond 3 weeks. We contacted Covid-19 online patient support groups (LongCovidSOS, Long Covid Support, Longcovid.org, Patient Safety Learning, Positive Path of Wellness, Patient Voices) from which we identified a third online group of UK doctors with long Covid), and also advertised on social media (Twitter) using the #longcovid hashtag. We used snowballing from our initial sample to identify further participants who were not in online groups or users of social media. Potential participants were screened for eligibility and baseline demographic information collected (Table 1). To correct for an initial gender and ethnic skew, we offered ‘men only’ focus groups and included additional rounds of invitations for participants from racial groups other than White British.

**Table 1:** Participant characteristics.

|  | Individual interviews | Focus group participants | Total |
| --- | --- | --- | --- |
| Total participants | 55 | 59 | 114 |
| Gender |  |  |  |
| • Female | 40 | 40 | 80 |
| • Male | 15 | 19 | 34 |
| • Other | 0 |  |  |
| • TOTAL | 55 | 59 | 114 |
| Age |  |  |  |
| • Median | 48 | 43 | 46 |
| • Range | 31-68 | 27-73 | 27-73 |
| Ethnicity |  |  |  |
| • White British | 39 | 45 | 84 |
| • White other | 4 | 4 | 8 |
| • Black | 3 | 2 | 5 |
| • Asian | 7 | 6 | 13 |
| • Mixed | 2 | 2 | 4 |
| Health professionals |  |  |  |
| • Doctor | 8 | 19 | 27 |
| • Nurse | 6 | 12 | 18 |
| • Paramedic | 1 |  | 1 |
| • Clinical psychologist | 1 |  | 1 |
| • Pharmacist | 1 |  | 1 |
| • Dental hygienist | 1 |  | 1 |
| • TOTAL | 1 |  | 1 |
| Recruited from |  |  |  |
| • Online patient group | 31 | 48 |  |
| • Social media | 17 | 11 |  |
| • Other | 7 |  |  |
| Hospitalised with acute Covid-19 |  |  |  |
| • Yes | 5 | 6 |  |
| • No | 50 | 53 |  |

[note to editor and reviewers: we are re-checking some of the figures in the above table – we know we had at least one physio for example – minor corrections will be made before final submission]

### Consent

All potential participants were sent brief details of the study and offered a more detailed standard information sheet. In accordance with ethics committee recommendations and infection control measures at the time (which discouraged exchange of paper documents), consent was collected either by email or verbally at the beginning of the audio or videotape. Participants were assured that all data would be de-identified and stored and handled anonymously, and that if they changed their mind about anything they said, they could contact a named researcher and withdraw that section of the data.

### Interviews

Interviewees were invited to tell their story uninterrupted and in their own words, with the interviewer using conversational prompts (such as “what happened next?” or “how did you feel when that happened?”) to maintain the narrative.^27^ Narrative interviews may be particularly useful for raising sensitive issues and identifying emotional touchpoints in an illness journey. All but one interviews were conducted by phone or by video using the business version of Zoom; one (of a participant who was not comfortable using video) was done by email at their preference. Phone and video interviews were audiotaped with consent and contemporaneous notes were also made. 11 individuals also submitted symptom diaries.

### Focus groups

Focus groups have the added advantage that group members may respond to the stories related by others, and group dynamics (e.g. humour, conflict) can be used as data, though lack of privacy may be a concern for some.^28^ Groups contained between 3 and 12 participants; they lasted 90 minutes (though participants were told they could leave any time if they were feeling tired). Each group had two trained facilitators including at least one clinically qualified researcher, plus a research assistant. Four groups were mixed; two were restricted to doctors, two to nurses and allied health professionals, and one to men (with male facilitators). One facilitator led the group; the other made contemporaneous notes and also captured real-time comments in the chat window, inviting comments on these as appropriate. In each group, all participants were asked to introduce themselves and outline briefly how Covid-19 had affected them before inviting positive stories about encounters with health services and then less positive experience.

### Data management and analysis

The first 10 individual interviews and all the focus groups were transcribed in full. For resource and speed reasons, and since many themes raised had already been covered, only selected portions of the other 45 interviews were transcribed. Transcripts and notes were de-identified by removing reference to real names and entered onto NVIVO software version 12. In an initial familiarisation phase, texts were grouped into broad categories (e.g. the illness experience, accessing services). An interim synthesis was produced from early transcripts and progressively refined using the constant comparative method (data from each new transcript were used to add nuance to the existing synthesis.^29^ Our analysis was informed by theories of illness experience,^30^ self-care,^30^ peer support,^31 32^ the clinical relationship,^33-35^ access to care,^32 36^ and service redesign^37^ (see Discussion).

### Patient involvement statement

The study was planned, undertaken, analysed and written in collaboration with people with long Covid. Two people with long Covid, both clinically qualified, assisted with the data interpretation and analysis and are listed as co-authors. We gave a webinar presentation via Zoom to which all 114 patient participants were invited (28 attended), and presented the key findings including the quotes used in this paper. A recording and copies of the presentation was shared with all participants and all were invited to correct any errors or misinterpretations. The draft paper was modified in response to their feedback.

## Results

### Description of dataset

Demographic and selected occupational details of participants are shown in Table 1. Despite our efforts to balance for gender and ethnicity, the final sample was skewed to 70% female and 75% White. By comparison, long Covid support groups are up to 86% female and the UK population is 80-85% White British (depending on how defined).^38^ The 55 interviews, 8 focus groups and 11 symptom diaries produced over 1000 pages of transcripts and notes. Our broad coding produced five over-arching themes which we discuss in more detail below: the illness experience, accessing care, relationships (or lack of) with clinicians, emotional touchpoints in encounters with health services, and ideas for improving services.

### A serious, uncertain and confusing illness

Participants described symptoms in every part of the body which were sometimes severe or fluctuating, made worse by the uncertain prognosis and stalled recovery, all of which combined to make this a frightening, confusing and debilitating illness. Many participants were unable to make sense of their suffering – an experience intensified by absence of medical knowledge or guidance. They described being trapped in a cycle of modest increments followed by setbacks which were physically and emotionally stressful, with no clear prospect of full recovery.

> I’ve been absolutely floored and I don’t know… I mean I’m just at the beginning of… I’ve got all sorts of… I’ve got… I see in [Town A] tomorrow, a rheumatologist, to find out what I’ve got because I’ve got vasculitis, which I think is a common thing […]. So, I don’t know how long that’ll unfortunately go on for. And I’ve been left with nerve issues, like really horrible nerve… stabbing pains in my hands and feet and I can’t move my toes anymore either, so I don’t know what the long term effects; I’m only at that point of just beginning to discover what the long term effects are, which are the ones that are kind of you expect to only affect people that were seriously ill in hospital, not the sort of the everyday people that managed at home with it really, so. So, I think my… unfortunately, my journey is far from over.
>
> (Individual interview WB1)

Many participants described themselves as previously fit and active, and as having had to come to terms with a dramatic decline in their ability to perform basic everyday activities.

> I’m trapped, in that I can’t park that far away and walk [to the shops] like I normally would because I can’t do hills. I can just, in the last couple of weeks, I can do gentle inclines now, but I sort of grind to a halt on a hill. So, it’s very limiting.
>
> (Individual interview TA1)

Participants had discovered the need to establish new routines and explicit self-disciplinary measures, such as counting steps and planning out visits to the shops, to avoid inevitable exhaustion if they over-exerted themselves:

> I find that it’s bizarre how… for instance, yesterday when I went to get my blood tests, I accidentally ended up getting five thousand steps on my step tracker because of the doctor’s trip and the pharmacy trip and, you know, moving around the house, getting dressed and stuff. It literally sent me crippled by six pm because it was just too much for me, compared to the days before where I’d done three thousand steps. So, even like the small increase like that just literally sends me completely washed out. (Individual interview EB1)

Symptoms such as fatigue and cognitive blunting (“brain fog”) severely limited the prospect of returning to work or finding new employment, as this office worker describes:

> I’m not working, I haven’t… I wasn’t able to go back to work and then I got made redundant. I’m… I can’t even imagine how I’m going to find a new job yet. In the last week, I’m wondering because my brain fog seems to have lifted and it’s feeling possible finally, after nearly six months, that I might one day find a new job. But my life is just nothing like it was and it’s not really the life I want, you know; I need to improve.
>
> (Individual interview TA1)

A few clinicians and non-clinicians in our sample made comparisons with post viral fatigue states like myalgic encephalomyelitis (ME) and chronic fatigue syndrome (CFS), expressing empathy and newfound solidarity with people suffering with these conditions. More commonly, our respondents felt the fatigue they were experiencing was distinct, and a consequence of their organ damage.

The level and duration of debility experienced by people in our sample often drew a negative response from friends, family or employer (as early expert advice suggested that non-hospitalized patients with mild disease would recover in two weeks^39^), especially when they had not had the disease confirmed by a test.

> The only reason I wanted the test is, however lovely friends are, it didn’t fit the two-week image they had of what this illness looks like. They said are you sure this isn’t anxiety? High pollen? It [wanting a positive test result] is more for validation. (Individual interview JH1)

Doctors and other clinicians in our sample described how their symptoms and the accompanying prognostic uncertainty, in addition to having all the effects listed above, had also stripped them of confidence in their professional abilities. They were especially afraid of the impact of cognitive deficits that might make them unsafe as clinicians. Some were keen to quantify their deficits by formal cognitive testing.

> [T]he medicolegal aspect is huge and I think possibly certainly feels that way as a GP and it’s scary to not be able to recognise potentially where you have deficits because if you can’t recognise them then that’s an unknown unknown in what can you do with that. And just the sort of fast-paced nature of GP and as [another participant] mentioned earlier you’ve got all these things having to kind of keep windows open at once. […] It seems terrifying to think how we’re going to get back to it
>
> (Participant from Doctors FG1)

In some cases, these professional concerns were compounded by the perception that clinical colleagues disbelieved or dismissed their symptoms as “just anxiety”.

### Difficulty accessing and navigating services

Participants found accessing care complex, difficult and exhausting. Many had contacted the national telephone advice service, NHS111, as directed by public messaging, and reported what they felt were delayed, absent or inappropriate responses due to pressure on the service itself, a perception among providers that their symptoms were less serious than they actually were, or lack of clearly defined care pathways for patients with long-term persisting symptoms.

> One day I had blue finger nails and I wasn’t cold and my husband was working at home at the time and I said to him and he looked, I mean I’d real proper cyanosis on all my finger nails and I phoned the GP and the GP answer phone said if you’ve got any of the signs of, of Covid please ring 111 and so I rang 111 and, I live in [city with high incidence of Covid-19] I don’t know if that makes any difference but I was put on hold and after over an hour, an hour and twenty minutes nobody answered so I just put the phone down because I was listening to music and a looped tape of what the symptoms were and I was getting, going crazy
>
> (Participant from Allied Health Professionals FG2)

Remote by default primary care services accessed through ‘total triage’ (which required every patient to complete an online consultation form or have a triage phone call) had been introduced as part of infection control policy in England and Wales in the acute phase of the pandemic.^40^ This system had generated additional queues and obstacles to getting seen by a general practitioner, which were particularly burdensome for participants whose disease was draining their energy or who did not own mobile phones.

> I’ve been able to get a lot of, book a lot of services online on the internet, they’ve now switched to Dr iQ which is only for mobile phones at the moment, I can’t afford my mobile phone, I have to phone in my deepest coma sleep at 8:00 am to talk to any GP now.
>
> (Participant in Patient FG1)

Some participants who had been discharged from hospital or contacted the national helpline NHS111 had been directed towards their general practitioner for managing long Covid symptoms (since the NHS111 route had been designed for acute Covid-19), but then directed back to NHS111. There was perceived to be a missing tier of support between patients self-managing their own symptoms and presenting acutely to hospital or being seen in a specialist clinic.

> The focus when you do get a new GP speaking to you seems to be that they go back to the beginning and I’ve had a few consultations where I know I don’t need to go to hospital but you’re assessment is really all around ‘do I stay at home and wait this out or do I go to hospital?’ and there’s nothing in between that. And I think if there was the same GP who we are able to consult regularly they would build a picture of your baseline and I think that’s what’s lost with digital ways of working.
>
> (Participant in Doctors FG2)

Some though not all general practitioners were reported to be unaware of rehabilitation services locally. Clinicians in our sample described finding out about rehabilitation clinics themselves and then asking for a referral. Few participants had been referred to a specialist and some who had received a specialist assessment described the experience as fragmented (they felt that “one bit of me” had been assessed by organ-specific tests and imaging, but there had no overall assessment of how long Covid had affected them generally and how they were functioning on a day to day basis). More often, they found local hospital outpatient services effectively closed for business, leaving them with no clear options.

> I’ve had to do a lot of this myself, to be honest. It was in the early on stages, I actually rang around the hospitals to see if there was anything, so, but there wasn’t anything. I just rang the switch board and said, ‘What’s the deal with people who’ve had Covid?’ But they said nothing. Gosh, yeah, I was desperate. I’m sorry, I’m one of these people who want answers and I wasn’t getting any answers’
>
> (Individual interview RH)

As the above quote illustrates, many participants did much work to self-advocate by emailing, telephoning or otherwise cajoling providers to make referrals or circumvent bottlenecks. These efforts occasionally included attempts – perhaps out of desperation – to ‘play’ an algorithm-driven system by omitting information (for example, deliberately conveying the impression to a receptionist or call handler that they had *not* had Covid-19). Others called on contacts or friends of friends to secure ‘back door’ appointments. Clinicians who used such tactics expressed guilt but also anger that most fellow sufferers lacked the kind of system knowledge that would allow them to do the same. Some non-clinicians, however, showed remarkable resourcefulness in this regard. The participant below describes her efforts to work around an NHS111 online form in order to be seen by her GP for a blood test:

> I did the e-consult – I had to do it a couple of times – I kind of learned to answer the questions to get it to send a message to my GP surgery… If you say you’ve got heart palpitations or breathlessness it’s telling you to call 111 which I didn’t want to do. And so I had to downplay symptoms [laughs] to get through. I cancelled it and did it again.
>
> (Patient AN1)

Another tactic employed, especially by clinicians who suspected they had end-organ damage, was to seek a private consultation. We heard several accounts of privately-ordered specialised tests (such as cardiac magnetic resonance imaging) which confirmed such damage; positive test results helped to validate their illness and (sometimes) gained them a specialist NHS referral.

### Concerns about quality and safety of care

Some of our participants described strong therapeutic relationships with particular clinicians, who listened to their stories, believed them, validated their suffering, acknowledged uncertainties, arranged appropriate tests and follow-up, and offered continuity of care.

> I was amazed at my experience because she was kind, empathetic she was compassionate [um] on the phone and she was listening to everything I was saying no matter how random or crazy the symptoms sounded but not only that she, I think for the first five days after I called her she had a daily check in call with me to monitor how I’m doing so it was like a ten minute phone call every day for the first five days (Participant in Patients FG1)

Unfortunately, such experiences were greatly outweighed in our dataset by the many who expressed concern that their clinician did not recognise their condition, did not believe that it existed, did not know how to diagnose it, did not empathise or acknowledge their suffering, did not know how to manage it (including ignorance of local rehabilitation services and referral pathways described above), and refused to test or refer. Some participants felt a responsibility, on behalf of fellow sufferers, to persuade clinicians that their symptoms were real, undertake their own research (individually or collectively) on this novel disease, and construct their own care pathway in the absence of an established local one. The following is from a patient who was getting troubling palpitations which they were aware could have a cardiac cause.

> They said ok we’ll get someone to phone you. My GP called back and just said ‘oh well it’s probably anxiety’. He didn’t seem to have any idea what it could be. I felt fobbed off. I said I’m worried – there are articles and news outlets that I’ve been reading and I want to know what’s happening to me – people are having strokes, blood clots. I haven’t been to hospital but I’m concerned I’m still getting these effects. He said ‘oh you’ll be fine you’ve only had it mildly’.
>
> (Individual interview ED1)

Whilst many participants liked the convenience of remote consulting (usually by telephone), others expressed concern about a lack of face-to-face examination, particularly for worrying symptoms. In some but by no means all cases, a request for a consultation was picked up by whoever was on duty, leading to loss of continuity of care – which mattered because the person’s story was often long, unusual and complex. Others described negative impacts on their clinical management (one patient with diabetes, for example, developed pain and tingling in the hands and feet and reduced ability to walk, and was surprised not to have been invited in to be examined).

Several participants related what may have been significant events which raised safety concerns. Some of these seemed to have been compounded by remote assessments and over-adherence to protocols.

> About five weeks in I think it was for me I was still desperately short of breath a little bit better than right at the start but it was still coming back in massive waves and I remembered ringing my GP from the floor on my lounge laying on my front and kind of saying I’m really short of breath, you know, do you think I should try an inhaler do I need to go back to A&E and I was kind of told well you don’t really sound too out of breath over the phone and I got given diazepam and I was just kind of just heartbroken at that point I was just absolutely like right I’m, I’ll take it so that I can tell you tomorrow maybe that that hasn’t helped or whatever but I just, I really felt at that point right if you could see me you would see that I am really like broken (Participant in Doctors FG2)

One interviewee described making a total of 16 phone calls to obtain a repeat prescription for an asthma inhaler. They had told a triage administrator that the long Covid had exacerbated their asthma, been told to contact NHS111 because this was what the protocol demanded for Covid-19 symptoms, but then been told by the NHS111 doctor to request the repeat prescription from their own doctor. The duty doctor eventually phoned them and told them they did not sound breathless enough to be given an inhaler. Further phone calls were needed to track down the regular general practitioner and secure the inhaler.

Given the many and evident gaps in services, many participants – both men and women – found that online peer support groups offered the greatest source of support through shared experiences, knowledge and validation. These groups contained considerable experiential expertise, and many participants heard about them from professionals who recognised the limitations of their own knowledge and understanding.

> At least I know I’m not alone. And I think people who actually have had the disease tend to know a little bit more about it. So, you know, sixth sense, I actually think that the support group has given more knowledge than the doctors have.
>
> (Patient EB1)

Discovery of such groups were described as “epiphany” moments and as “salvation” by participants, giving them hope that they might begin to move on from the chaos of long Covid illness and onto a story of restitution.^41^

### Emotional touch points

The experiences described above were associated with what Bate and Robert have called emotional touch points:^42^ powerful feelings such as anger, frustration, fear and hopelessness. Our participants described feeling physically and emotionally exhausted from the burden of trying to access services, be believed, navigate incomplete and inadequate care pathways, gain knowledge, organise their own recovery plan, and integrate their own care across a disjointed and siloed service. Some talked of a profound breakdown of trust in a previously valued family doctor service.

When asked what changes would be needed in services to avoid or repair these emotional touch points, participants made many suggestions, from which we distilled some preliminary quality principles for a long Covid service (see Box 1).

**Box 1: Quality principles for Long Covid services**

1. **Access** Everyone with long Covid should have access to appropriate care, whether or not they have had a positive laboratory test for Covid-19 or a hospital admission.
2. **Burden of illness** The burden on the patient for accessing, navigating and coordinating their own care should be minimised. Care pathways should be clear and referral criteria explicit.
3. **Clinical responsibility and continuity of care** Clinical responsibility for the patient should be clear. Whilst specialist investigation and management of particular complications is important, one clinician should take care of the whole patient and provide continuity of care.
4. **Multi-Disciplinary rehabilitation services** Patients requiring a formal rehabilitation package should be assessed by a multi-disciplinary team including (e.g.) rehabilitation, respiratory and cardiac consultant, physiotherapist, occupational therapist, psychologist and (if needed) neurologist.
5. **Evidence-based standards** Standards and protocols should be developed, published and used so that investigation and management is consistent wherever care is received.
6. **Further development of the knowledge base and clinical services** Clinical teams should proactively collect and analyse data on this new disease so as to improve services and build the knowledge base. Patients should be partners in this endeavour. As a first step, patients need to be counted and prevalence rates and prognosis established.

## Discussion

### Summary of key findings

This qualitative study of 114 people with long Covid in UK, including high representation from health professionals, has revealed a number of important findings. People experience long Covid as a confusing illness with many, varied and often relapsing-remitting symptoms, uncertain prognosis and a heavy sense of loss and stigma. They find it difficult to access and navigate services which they experience as fragmented and siloed; some described not being taken seriously. There appears to be wide variation in clinical practice (e.g. inconsistent criteria for seeing, investigating and referring patients), and in the quality of the therapeutic relationship. We identified a number of possible critical events which may have been partly due to overstretched, disjointed services designed to discourage face-to-face encounters. These findings informed draft quality principles, summarised in Box 1.

### Comparison with theoretical literature

Unlike disease, which can be defined in terms of a typical constellation of symptoms, signs and test results, illness is a personal, lived experience that is both emotionally laden and socially meaningful.^30^ Participants’ experiences of Covid-19 and its sequelae fitted Frank’s depiction of the “chaos narrative”, in which an illness experience is uncertain, confusing and with no clear direction or purpose.^41^ Many of the narratives conveyed a sense of shame and blame consistent with stigma.^43^ Our findings illustrate that serious illness does not merely disrupt people’s activities and routines; it can threaten their very identity as healthy, independent and successful selves.^44 45^ “Spoiled identity” seemed a particular concern in our sample, especially since symptoms were largely non-specific and not biomedically validated.

The heavy burden of treatment described by our participants, where the patient is expected to self-manage some or all aspects of their care, accords with contemporary theories of illness burden.^46^ Accounts of positive care experiences are explained by theories of good professional practice.^35^ The good clinician engages reflexively with the patient’s lived experience and acts not merely as diagnostician or technical expert but also as active listener and professional witness.^33 34^ Continuity of the clinical relationship is particularly important in chronic illness.^34^ These qualities are enshrined within the professional standards outlined in the General Medical Council’s ‘Duties of a Doctor’.^47^

Our data are consistent with previous theorisations of access to healthcare in relation to objectively defined dimensions of the service (e.g. approachability, acceptability, availability, affordability, appropriateness^48^). Patients’ struggles to be seen as legitimate and gain access to services also resonate with the sociological notion of *candidacy* – that is, how the patient’s eligibility for care is negotiated between them and their healthcare providers (a construct which is structurally, culturally, organisationally and professionally shaped).^49 50^

Making sense of, and managing, chronic illness may become easier in peer support communities (often though not always online), where new members learn practical approaches from more experienced ones and that was certainly the case amongst our cohort, who frequently felt this was their only option.^32 36^

The identification of emotional touch-points in our participants’ narratives draws on experience-based [co-]design (EBCD), an approach developed to ensure that health services are designed around the patient experience.^37^ Key features include a grounding in the perceptions and reactions of individual patients, pragmatic application in front-line health services, and the use of collective sensemaking to produce new understandings.^37^

### Strengths and limitations of the study

To our knowledge, this is the largest and most in-depth qualitative study of long Covid published in the academic literature to date. The research team included both clinicians and social scientists. Our participants spanned a wide range of ages, ethnic and social backgrounds, and illness experiences – including, importantly, the under-researched majority who were never hospitalised. We oversampled men and people from non-White ethnic groups to partially correct an initially skewed sample. Offering the choice of interviews or focus groups allowed those with sensitive stories to tell to do so in private, and for all participants to select a method with which they were comfortable. The use of multiple linked sociological theories allowed to produce a rich theorisation of the lived experience of the illness and draw on that theorisation to produce principles and practical proposals for improving services. We included experts by experience (people with long Covid) as steering group members, co-interpreters of the data, co-authors on the paper and peer reviewers. The inclusion of a high proportion of healthcare workers both reflects the occupational risk in these groups^51 52^ and allowed participants to draw on their system knowledge as well as their personal illness experience when suggesting improvements to services.

The study does, however, have some limitations. The sample was drawn entirely from the UK, though we hope to go on to produce cross-national comparisons by collaborating with researchers in other countries. We did not fully correct for skews in the sample, and in particular the perspectives of some minority ethnic groups have not been fully captured. It is likely that despite our efforts to do democratic collaborative research *with* patients, we may not have fully grasped the lived experience or represented all voices.

### Comparison with previous empirical studies

Perhaps the most important contribution our findings make to previously published data is to affirm the experience of patients with long Covid previously published in auto-ethnographies,^53^ patient-led surveys,^4^ narrative reviews,^54^ commentaries,^1 55^ and the views of ‘doctors as patients’.^56^ Our findings also resonate to a large extent with surveys on patients discharged from hospital.^7 9 57^ These sources, like our own dataset, emphasise the varied manifestations, uncertain course and sometimes protracted recovery from this disorienting and new illness; the stigma of not being believed; the frustration of scientific and medical uncertainty and ignorance; and problems accessing health services. We have captured the voices of patients who were excluded from other formal research studies because they were not admitted to hospital. We have taken forward suggestions for new services and improvements in healthcare, based on the emotional touch points of the patient experience.

### Implications for services

A wholescale redesign of clinical services for long Covid is beyond the scope of this paper. Moreover, the details of such services will be determined by local and contextual factors. However, based on our data, we believe that the quality principles listed in Box 1 should inform and underpin both generalist and specialist services for long Covid.

Many of these principles accord strongly with a recently published manifesto written by a group of doctors with long Covid which calls for: greater emphasis on research and surveillance; appropriate development of, and access to, clinical services; and significant patient involvement in both research and service development.^56^

Whilst policymakers in UK have recently announced a new service for Covid-19 rehabilitation in specialist clinics, ^25^ primary care services are not mentioned. Based on our findings, general practitioners and other primary care clinicians appear to need better knowledge, better guidance, and more time and resources to deliver the generalist care and support which many patients with long Covid need, though this would have resource implications.

## Conclusion

This study has illustrated the uniquely varied, burdensome and uncertain nature of the lived experience of long Covid and provided some preliminary principles for developing services to address their needs (Box 1).

Perhaps the most important contribution of this study is to have identified a mismatch between what the peer-reviewed scientific literature is saying about persistent symptoms and what patients are actually experiencing. Our findings suggest that the various online patient support groups for long Covid have earnt a place at the table when designing services and planning new research. Whilst such groups are not new,^31^ and may sometimes grow to a membership of thousands and provide a source of patient-generated data for researchers,^58^ the long Covid peer support groups on Facebook and Slack have taken the lead in generating the evidence base on long Covid,^4^ sharing practical advice, filling gaps in the formal support system, and campaigning for better and more consistent services.

While health social movements studied by sociologists have traditionally been founded and driven by non-clinically trained patients,^36^ long Covid is perhaps novel in including relatively large numbers of clinicians and scientists among its ranks (many of whom contracted the virus in the course of their work and now have multisystem complications). The combination of clinical *and* experiential knowledge offered by such communities is an important resource for both service planning and research.^56^ It is time to collaborate with these groups to take forward a genuinely patient-centred research agenda.

## Data Availability

Anonymised data from this qualitative study can be made available to other qualitative researchers on discussion with the authors.

## Acknowledgements

*We thank the 114 participants for their interest and contributions, and xx peer reviewers for helpful comments on a previous draft of this paper*.

## Contributors and sources

EL, AR and TG conceptualised and designed the study. EL, AR, SW and TG conducted interviews and focus groups. EL and AR led data analysis, with input from SW and TG, and produced a first draft of the results section. TG wrote the first draft of the paper which was refined by all authors. LH provided research assistant support and conducted some interviews. ST and CR provided expertise by experience and knowledge of patient-led research. TG presented findings to long Covid patient participants with assistance from EL, AR, SW and LH. All authors contributed to refinement of the paper provided additional references. TG is corresponding author and guarantor. TG affirms that the manuscript is an honest, accurate, and transparent account of the study being reported; that no important aspects of the study have been omitted; and that any discrepancies from the study as planned (and, if relevant, registered) have been explained.

## Copyright

The Corresponding Author has the right to grant on behalf of all authors and does grant on behalf of all authors, a worldwide licence to the Publishers and its licensees in perpetuity, in all forms, formats and media (whether known now or created in the future), to i) publish, reproduce, distribute, display and store the Contribution, ii) translate the Contribution into other languages, create adaptations, reprints, include within collections and create summaries, extracts and/or, abstracts of the Contribution, iii) create any other derivative work(s) based on the Contribution, iv) to exploit all subsidiary rights in the Contribution, v) the inclusion of electronic links from the Contribution to third party material where-ever it may be located; and, vi) licence any third party to do any or all of the above.

